# Tobacco use and determinants among adults with non-communicable diseases: Evidence from the 2017 Zambia STEPS survey

**DOI:** 10.64898/2026.05.15.26353278

**Authors:** Chola Bwalya, Given Moonga, Mwiinde M. Allan, Carla Berg, Adam Silumbwe, Cosmas Zyambo

## Abstract

**Background:** Non-communicable diseases (NCDs) account for approximately 75% of global deaths, with 79% occurring in low- and middle-income countries. Tobacco use remains a major modifiable risk factor, contributing to more than 8 million deaths annually. In Zambia, evidence on tobacco use among individuals with hypertension, diabetes mellitus, and cardiovascular disease remains limited. This study assessed the prevalence and determinants of tobacco use among adults with NCDs in Zambia.

**Methods:** We conducted a secondary analysis of the 2017 Zambia STEPS survey. The analytic sample included 716 adults aged 18–69 years with self-reported hypertension, diabetes, and/or cardiovascular disease. Tobacco use was defined as current smoking or smokeless tobacco use. Multivariable logistic regression was used to estimate adjusted odds ratios (AORs), accounting for the complex survey design.

**Results:** Among 716 participants, 65.5% had hypertension, 7.7% diabetes, and 26.8% cardiovascular disease; 89.5% had multimorbidity. The overall prevalence of tobacco use was 12.2%. Prevalence was 12.2% among those with hypertension, 5.5% among those with diabetes, and 14.1% among those with cardiovascular disease. Tobacco use was significantly higher among males. Female sex was associated with lower odds of tobacco use (AOR = 0.16, 95% CI: 0.05–0.54, p = 0.004). Secondary education (AOR = 0.15, 95% CI: 0.03–0.66) and higher education (AOR = 0.04, 95% CI: 0.01–0.44) were protective. Alcohol consumption increased the odds of tobacco use (AOR = 5.23, 95% CI: 1.17–23.28).

**Conclusion:** Tobacco use remains common among adults with NCDs in Zambia. Integration of tobacco cessation interventions into routine NCD care is urgently needed.

## Introduction

Despite global declines in tobacco use, smoking continues to be the leading preventable health risk worldwide [1][2]. The World Health Organization (WHO) estimates over seven million annual tobacco-related deaths[3] positioning it as a major risk factor for non-communicable diseases (NCDs)[4]. Tobacco kills up to half of those who use tobacco and do not quit[3], contributing substantially to the global burden of premature mortality[4]. By 2030, tobacco is projected to claim over 8 million lives annually, with more than 80% deaths in low and middle-income countries[5].

NCDs account for a substantial proportion of global mortality, with at least 43 million deaths in 2021, representing 75% of non-pandemic related deaths worldwide [4]. Cardiovascular diseases (CVD) and diabetes mellitus (DM) are among the most fatal, claiming 17.9 million and 1.6 million deaths annually, respectively [6] Hypertension (HTN) a significant risk factor for CVD, affects approximately 1.13 billion people globally[7].

In Sub-Saharan Africa (SSA), the burden of NCDs has escalated significantly, with disability-adjusted life years (DALYs) attributable to NCDs increasing by 67% between 1990 and 2017, largely driven by CVD [8]. Despite well-documented risks, evidence shows that individuals living with NCDs continue tobacco use [9][10][11]. In recognition of the global epidemic, WHO member states adopted the Framework Convention on Tobacco Control (FCTC) to guide comprehensive interventions aimed at reducing tobacco use [3].

Tobacco related illnesses in Zambia contribute to approximately 7,900 deaths annually [12], underscoring the urgent need for effective control measures. Although Zambia ratified the WHO FCTC in 2008, and implemented interventions such as bans on tobacco advertising, restrictions on smoking in educational and healthcare facilities, and limitations on access to free tobacco products [13]; tobacco use has continued to rise, this increase is driven by low cigarette prices, which remain 13.5% more affordable than the African average [14]. As of 2016 tobacco taxes accounted for only 37% of retail prices, far below recommended 75% threshold [15]. Furthermore, a five year tax holiday aimed at attracting domestic manufacturing was extended to the tobacco industry, resulting in the establishment of new production facilities and increased cigarette output[16].

Despite several studies examining factors associated with tobacco use in Zambia[13][17][18][15][19], there remains limited evidence on tobacco use among adults with NCDs. This gap in knowledge may hinder the design of targeted control strategies for high risk groups and represents missed opportunity, to integrate cessation support into NCD care. Such integration could reduce complications, mortality, and healthcare costs associated with managing NCDs.

This study therefore aims to assess the factors associated with tobacco use among adults 18-69 years with HTN, DM, and CVD in Zambia. By identifying factors associated with tobacco use in this population, the findings will provide critical insights to inform tailored interventions and strengthens tobacco control policies within NCD care frameworks.

## Materials and methods

### Study design and data source

This study involved secondary data analysis. The analytic sample for the current study was adults aged 18-69, diagnosed with HTN, DM, and CVD. This cross-sectional design provides a snapshot of the population at a specific point in time allowing for analysis of associations between variables. The Zambia STEP-wise survey for NCD prevalence and risk factors, mental and oral health conducted in 2017 was a cross-sectional national survey designed to obtain data representative of the adult population, ages 18 to 69, living in Zambia [20]. The STEPS survey used a multi-stage cluster sampling technique and utilized the household listing from the Zambia Population-Based HIV Impact Assessment (ZAMPHIA). The first stage of sampling involved selection of Standard Enumeration Areas (SEAs) from each of the 10 provinces using probability proportional to size (PPS); in the second stage, 15 households in each of the rural SEAs and 20 households in each of the urban SEAs were selected systematically using appropriate sampling intervals based on the number of households in that SEA. Thirdly, one member of the household who was eligible was purposively selected for an interview. The questionnaire administered to survey participants included demographic data, as well as physical and biochemical measurements. Overall 4, 301 participants were included in the survey, with a response rate of 77.7%. Details of the WHO STEPS methodology can be found elsewhere [13][15].

### Study participants

The analytic sample for the current study was adults ages 18-69, diagnosed with HTN, DM, and CVD including those who are currently use tobacco (smoking or smokeless tobacco), those who formerly used tobacco (quit at least 12 months before participating in the study), and those who have never used tobacco (never used tobacco), all of whom consented and were enrolled during the time of data collection. Participants with missing data such as diagnosis, tobacco use status and demographic information needed for analysis were excluded from the study. In the STEPS dataset (n=4,903), 3,586 respondents did not report an NCD diagnosis and were excluded, leaving 716 eligible participants. Within this subgroup although the exclusion criteria specified removal of participants with missing data on diagnosis, tobacco use status, or key demographic variables, all 716 respondents included in the analytic sample had complete data on these variables of interest. Therefore, no participants were excluded on the basis of missing data.

### Variables and statistical analysis

#### Primary dependent variables

The primary dependent variable was tobacco use. We defined tobacco use as current daily or non-daily use of combustible and non-combustible tobacco products [21]. Participants were asked: 1) “Do you currently smoke any tobacco products, such as cigarettes, shisha, cigars, or pipes?” and 2) “Do you currently use any smokeless tobacco products such as snuff chewing?” Participants answering YES to either were categorized as people who use tobacco; those answering NO to both were categorized people who do not use tobacco.

#### Independent variables

Independent variables were chosen based on the literature regarding potential predictors for tobacco use including: sociodemographic characteristics, alcohol consumption, tobacco-related exposures, and NCD status.

##### Sociodemographic variables

The independent variables included the following socio-demographic variables: age, gender (male, female), residence (rural, urban), education (no formal schooling, primary school, secondary school, and higher education), marital status (married not married), and employment (not employed, employed), and income.

##### Behavioral variables

The participants were asked, “Have you consumed any alcohol within the past 12 months?” (Categorized as yes vs. no past-year alcohol use).

##### Environmental and media exposure variables

Participants were asked: 1) “During the past 30 days did someone smoke in your home?” (Categorized as yes vs. no past-30-day home exposure); 2) “During the past 30 days did someone smoke in closed areas in your work place (in the building, a work area or a specific office)?” (Categorized as yes vs. no past-30-day workplace exposure); 3) “During the past 30 days, have you noticed any of the following types of cigarette promotions? Free sample gifts, cigarettes at reduced prices, free gifts or special discounts on buying cigarettes, clothing items or other items with a cigarette brand name or logo, cigarette promotions in the mall” (categorized as any vs. no tobacco promotion exposure); And 4) “During the past 30 days have you noticed any information about the dangers of smoking cigarettes or that encourages quitting through the media? Radio?” (Categorized as any vs. no tobacco prevention message exposure).

##### NCD status

Participants were asked: 1) “Have you ever been told by a doctor or other health worker that you have: a raised blood sugar or diabetes?” (Categorized as yes vs. no DM diagnosis); 2) “Have you ever been told by a doctor or other health worker that you have a raised blood pressure or hypertension?” (categorized as yes vs. no HTN diagnosis); and 3) “Have you ever had: heart attack, stroke, currently using ASA to prevent or treat heart disease and currently taking statins regularly to prevent or treat heart disease?” (Those who answered yes to any of these questions were categorized as having CVD; others were categorized as no CVD diagnosis).

### Statistical Analysis

Weighted analysis for complex surveys was conducted, including descriptive analysis, which was used to summarise the characteristics of the study population and to estimate the prevalence of tobacco use among adults aged 18–69 years with HTN, DM, and CVD. The continuous variable age was assessed for normality using the Shapiro–Wilk test and was found to be skewed; therefore, results are presented as medians and interquartile ranges (IQRs). The Mann–Whitney U test (Wilcoxon rank-sum test) was used to assess associations between tobacco use and continuous variables, while the chi-square test was used to examine associations between tobacco use and categorical variables. Frequencies and percentages are reported.

Inferential analysis focused on tobacco use as a binary outcome, with logistic regression employed to identify associated factors. All analyses accounted for the complex survey design by incorporating sampling weights, primary sampling units (PSUs), and strata to ensure population-representative estimates and valid standard errors. Sampling weights (popwt3) adjusted for unequal selection probabilities and non-response, while PSUs were defined as enumeration areas selected in the first stage of the STEPS survey, and strata were based on province and urban/rural residence. The survey design was declared in Stata using the command svyset psu [pweight = popwt3], strata (stratum), thereby incorporating clustering, stratification, and weighting. Analyses were conducted using Stata’s *svy* suite of commands, which apply design-based weights and variance estimation methods to produce nationally representative estimates. Logistic regression models were fitted within the generalized linear model (GLM) framework, with both univariable and multivariable analyses performed to assess crude and adjusted associations between explanatory variables and tobacco use.

### Ethical approval

The current study, based on secondary data, was reviewed and approved by the University of Zambia Biomedical Research Ethics Committee (UNZABREC, Ref. No. 6283-2025). The STEPS Survey had prior approval from UNZABREC, and written informed consent was obtained from all participants [20].

## Results

Table 1 presents the prevalence of tobacco use among adults with NCDs. Overall prevalence was 12.2%, higher in men (22.5%) than women (8.6%). By condition, prevalence was 12.2% in HTN (57 participants), 5.5% in DM (3 participants), and 14.1% in CVD (27 participants). Tobacco use was most common among males, rural residents, and those without formal education, with a median age of 50 years (IQR: 34–60).

**Table 1.**
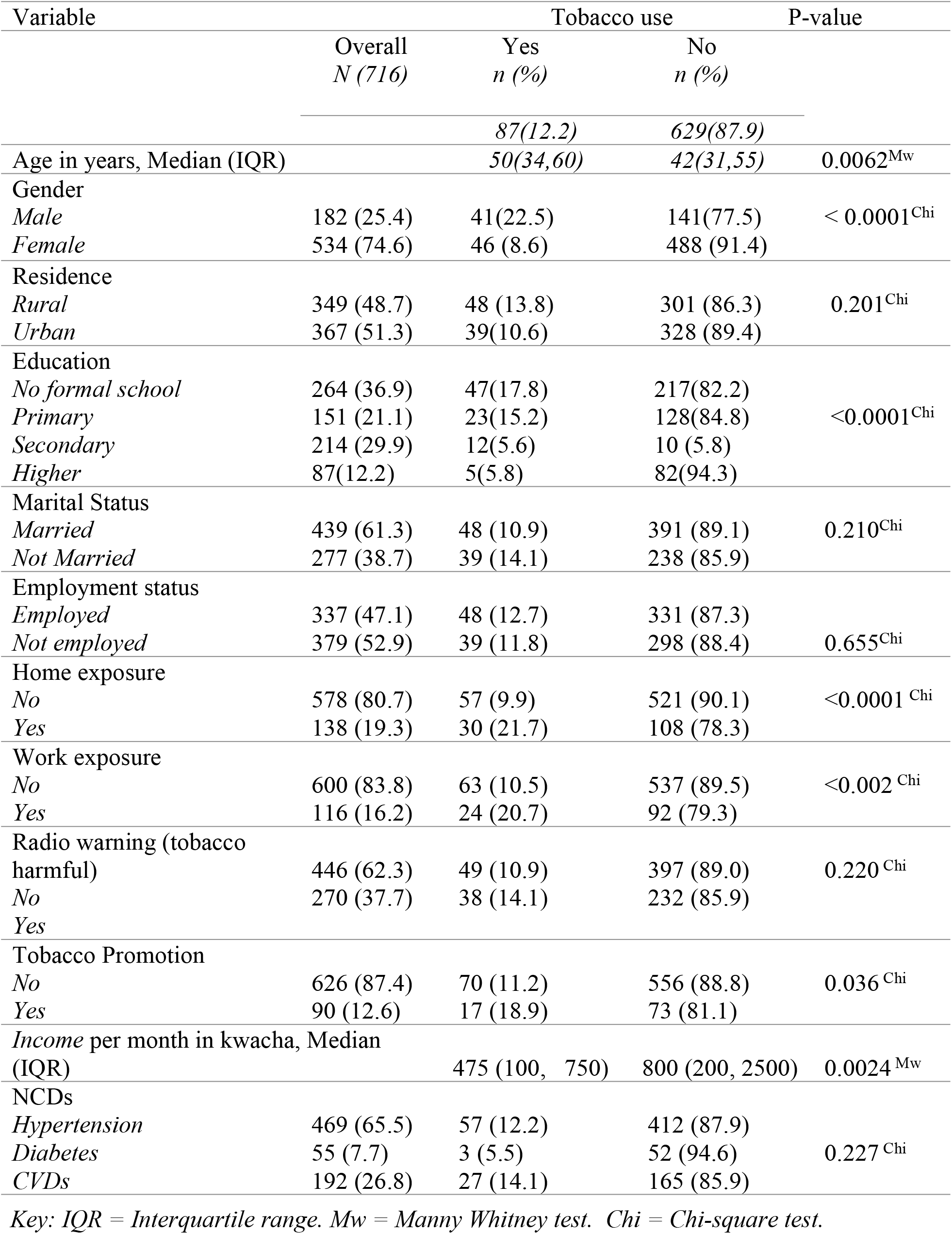
Distribution of sociodemographic factors and associated P-values among adults with hypertension, diabetes, and cardiovascular disease by tobacco use status in Zambia.

### Factors associated with tobacco use

Table 2 presents factors associated with tobacco use among adults with NCDs in Zambia. Univariate logistic regression identified, age, gender, residence, education, home exposure, work exposure, tobacco promotion as significant factors. Multivariable analysis provided adjusted estimates: women had significantly lower odds of using tobacco compared to men (AOR = 0.16, 95% CI: 0.05–0.54, p = 0.004). Compared to individuals with no formal schooling, having secondary education was associated with reduced odds of tobacco use (AOR = 0.15, 95% CI: 0.03– 0.66, p = 0.013), while higher education further reduced the odds (AOR = 0.04, 95% CI: 0.01– 0.44, p = 0.008). In contrast, alcohol consumption had substantially higher odds of using tobacco (AOR = 5.23, 95% CI: 1.17–23.28, p = 0.031).

**Table 2.** Logistic regression estimates for factors associated with tobacco use among adults with hypertension, diabetes, and cardiovascular disease in Zambia.

| Variable | UOR 95% CI | P-value | AOR 95% CI | P-value |
| --- | --- | --- | --- | --- |
| Age in years | 1.02 (1.00-1.04) | 0.017 | 1.01 (0.97-1.05) | 0.544 |
| Gender |  |  |  |  |
| *Male* | ref |  | ref |  |
| *Female* | 0.32 (0.19-0.52) | <0.0001 | 0.16 (0.05-0.54) | 0.004 |
| Residence |  |  |  |  |
| *Rural* | ref |  | ref |  |
| *Urban* | 0.68 (0.43-1.90) | 0.113 | 0.45 (0.13-1.56) | 0.207 |
| Education |  |  |  |  |
| *No formal school* | ref |  | ref |  |
| *Primary* | 0.80 (0.45-1.43) | 0.446 | 0.78 (0.19-3.27) | 0.733 |
| *Secondary* | 0.26 (0.13-0.52) | <0.0001 | 0.15(0.3-0.66) | 0.013 |
| *Higher* | 0.34 (0.14-0.83) | 0.018 | 0.04(0.01-0.44) | 0.008 |
| Marital status |  |  |  |  |
| *Married* | ref |  | ref |  |
| *Not Married* | 1.45 (0.90-2.34) | 0.131 | 1.11 (0.33-3.77) | 0.864 |
| Employment |  |  |  |  |
| *Employment* | ref |  | ref |  |
| *Not employed* | 0.83 (0.53-1.29) | 0.403 | 1.07 (0.36-3.12) | 0.904 |
| Alcohol consumer |  |  |  |  |
| *No* | ref |  | ref |  |
| *Yes* | 3.48 (1.65-7.30) | 0.001 | 5.23 (1.17-23.28) | 0.031 |
| Home exposure to tobacco |  |  |  |  |
| *No* | ref |  | ref |  |
| *Yes* | 2.44 (1.47-4.04) | 0.001 | 1.34 (0.40-4.43) | 0.631 |
| Work exposure to tobacco |  |  |  |  |
| *No* | ref |  | ref |  |
| *Yes* | 2.23 (1.26-3.93) | 0.006 | 0.41 (0.10-1.71) | 0.219 |
| Exposed to tobacco promotion |  |  |  |  |
| *No* | ref |  | ref |  |
| *Yes* | 1.74 (0.99-3.06) | 0.055 | 4.62 (0.82-25.9) | 0.082 |
| Radio warning (tobacco harmful) |  |  |  |  |
| *No* | ref |  | ref |  |
| *Yes* | 1.29 (0.81-2.06) | 0.274 | 0.52 (0.17-1.64) | 0.263 |
| Income per month in Kwacha | 0.99 (0.99-1.00) | 0.413 | 0.99 (0.99-1.00) | 0.222 |
| Key: OR=Odds Ratio, UOR=Unadjusted Odds Ratio, AOR=Adjusted Odds Ratio, CI=95% confidence interval, ref=reference category |  |  |  |  |

## Discussion

This study assessed the prevalence and determinants of tobacco use among adults living with non-communicable diseases (NCDs) in Zambia using nationally representative STEPS survey data. The findings show that tobacco use remains a significant behavioural risk factor among individuals already diagnosed with hypertension, diabetes mellitus, or cardiovascular disease. The overall prevalence of tobacco use (12.2%) highlights the persistence of tobacco consumption even among high-risk clinical populations, underscoring missed opportunities for cessation interventions within NCD care pathways.

The observed prevalence among adults with NCDs is comparable to estimates from other studies in Zambia and the broader region, although variation exists depending on population groups and study design. Similar levels have been reported among hypertensive patients in Africa, while higher estimates have been observed in clinical populations in other low- and middle-income countries. These findings reinforce the fact that tobacco use remains common among individuals with chronic diseases despite known risks of disease progression, complications, and premature mortality. From a clinical perspective, continued tobacco use after diagnosis suggests gaps in integration of cessation counselling within routine NCD management services.

A key finding from this study is the strong protective association between female sex and tobacco use. Women were significantly less likely to use tobacco compared to men, consistent with evidence from Zambia and other low- and middle-income settings. This gender disparity is widely documented and is largely attributed to social norms, cultural expectations, and gendered patterns of substance use. In many settings, tobacco use is socially more acceptable among men than women, which contributes to sustained higher prevalence in males. However, the persistence of tobacco use among women, although lower, remains an important public health concern given emerging evidence of increasing uptake in some populations. These findings suggest that tobacco control interventions should remain gender-sensitive while also monitoring changing behavioural trends among women.

Educational attainment demonstrated a clear and consistent protective gradient against tobacco use. Individuals with secondary and higher education were significantly less likely to use tobacco compared to those with no formal education. This finding is consistent with a large body of evidence indicating that education influences health behaviour through multiple pathways, including improved health literacy, better access to health information, and enhanced capacity for informed decision-making. Higher education may also be associated with increased awareness of the health risks of tobacco use and greater exposure to cessation messaging. From a policy perspective, these findings highlight the importance of strengthening health education and communication strategies targeted at populations with low educational attainment.

Alcohol consumption was strongly associated with tobacco use, with individuals who consumed alcohol being more than five times more likely to use tobacco. This co-occurrence of alcohol and tobacco use has been widely documented and reflects shared behavioural, psychological, and social determinants. One explanation is the behavioural clustering of risk factors, where individuals engaged in one risky behaviour are more likely to engage in others. Additionally, both substances may be used as coping mechanisms for stress or social pressures, as suggested by behavioural and psychosocial theories of substance use. The strong association observed in this study underscores the need for integrated interventions addressing multiple risk behaviours rather than isolated tobacco control strategies.

In contrast, environmental exposure variables such as home and workplace smoke exposure, as well as exposure to tobacco promotion, were not independently associated with tobacco use after adjustment. Although these factors showed significant crude associations, their effects were attenuated in multivariable analysis, suggesting confounding by sociodemographic and behavioural factors. This may indicate that environmental exposures operate indirectly or are closely correlated with underlying behavioural and socioeconomic determinants. It may also reflect limitations in measurement using self-reported exposure variables, which are subject to recall and reporting bias.

Importantly, this study highlights that tobacco use among individuals with NCDs is shaped by a combination of individual-level and behavioural determinants rather than environmental exposure alone. The findings align with the socio-ecological model, which conceptualises health behaviours as being influenced by interactions between individual characteristics, social relationships, and broader environmental and policy contexts. In this case, gender, education, and alcohol use operate at the individual and behavioural levels, while tobacco exposure variables reflect household and workplace environments that may reinforce use but are not sufficient determinants on their own. From a health systems perspective, the persistence of tobacco use among individuals already diagnosed with chronic diseases represents a critical gap in secondary prevention. NCD clinics provide an important opportunity for integrating tobacco cessation interventions into routine care. However, the findings suggest that such integration may currently be inadequate or inconsistently implemented. Strengthening brief cessation counselling, improving access to cessation support tools, and embedding behavioural risk screening into NCD management protocols may improve outcomes in this population.

### Strengths and limitations

This study has several strengths. It used nationally representative STEPS survey data, allowing for generalisable findings across Zambia. The use of survey weights and adjustment for complex sampling design strengthens the validity of the estimates. Additionally, focusing specifically on individuals with diagnosed NCDs provides clinically relevant insights for targeted interventions. However, the study also has limitations. The cross-sectional design limits causal inference between identified determinants and tobacco use. Self-reported measures may introduce social desirability and recall bias, potentially leading to underestimation of tobacco use. Additionally, the analysis does not capture temporal changes in behaviour following NCD diagnosis, which would be important for understanding cessation dynamics.

## Conclusion

Tobacco use remains a significant behavioural risk factor among adults living with NCDs in Zambia. The findings demonstrate that tobacco use is primarily driven by gender, education, and alcohol consumption, reflecting a complex interplay of social and behavioural determinants. These results highlight the need for integrated, multi-level interventions that combine clinical cessation support within NCD services with broader population-level prevention strategies.

## Data Availability

The data is available together with submission of this manuscript

## Acknowledgements

We gratefully acknowledge the WHO NCD Microdata Repository for granting access to the data used in this study. We express our profound gratitude to Professor Choolwe Jacobs, Mr. Danny Kabwe, and Mr. Nathan Tembo, for their invaluable contributions to the study.

